# Urinary metabolite profiling identifies biomarkers for risk of progression of diabetic nephropathy in 2,670 individuals with type 1 diabetes

**DOI:** 10.1101/2020.10.21.20215921

**Authors:** Stefan Mutter, Erkka Valo, Viljami Aittomäki, Kristian Nybo, Lassi Raivonen, Lena M Thorn, Carol Forsblom, Niina Sandholm, Peter Würtz, Per-Henrik Groop

**Affiliations:** Folkhälsan Institute of Genetics, Folkhälsan Research Center, Helsinki, Finland; Abdominal Center, Nephrology, University of Helsinki and Helsinki University Hospital, Helsinki, Finland; Research Program for Clinical and Molecular Metabolism, Faculty of Medicine, University of Helsinki, Helsinki, Finland; Nightingale Health Ltd, Helsinki, Finland; Department of General Practice and Primary Health Care, University of Helsinki and Helsinki University Hospital, Helsinki, Finland; Department of Diabetes, Central Clinical School, Monash University, Melbourne, Victoria, Australia

**Author notes:** Corresponding author: Professor Per-Henrik Groop, Folkhälsan Research Center, Biomedicum Helsinki, Haartmaninkatu 8, FIN-00290 Helsinki, Finland.

**Keywords:** Diabetic nephropathy, progression, NMR, metabolite profiling, type 1 diabetes

## Abstract

**Aims:** This study examines associations between 51 urinary metabolites and risk of progression of diabetic nephropathy in individuals with type 1 diabetes by employing an automated nuclear magnetic resonance (NMR) metabolomics technique suitable for large-scale urine sample collections.

**Methods:** For 2,670 individuals with type 1 diabetes from the FinnDiane Study, we collected 24-hour urine samples and measured metabolite concentrations by NMR. Individuals were followed for 9.0 ± 5.0 years until their first sign of progression of diabetic nephropathy, end-stage kidney disease or study end. Cox regression analyses were performed on the entire study population (overall progression), on 1,999 individuals with normoalbuminuria and 347 individuals with macroalbuminuria at baseline.

**Results:** Seven urinary metabolites were associated with overall progression after adjustment for baseline albuminuria and chronic kidney disease stage (p < 8 × 10^-4^): Leucine (hazard ratio 1.47, 95% confidence interval [1.30, 1.66] per 1-SD creatinine-scaled metabolite concentration), valine (1.38 [1.22, 1.56]), isoleucine (1.33 [1.18, 1.50]), pseudouridine (1.25 [1.11, 1.42]), threonine (1.27 [1.11, 1.46]) and citrate (0.84 [0.75, 0.93]). 2-hydroxyisobutyrate was associated with overall (1.30 [1.16, 1.45]) and also progression from normoalbuminuria (1.56 [1.25, 1.95]). Six amino acids and pyroglutamate were associated with progression from macroalbuminuria.

**Conclusions:** Branched-amino acids and other urinary metabolites were associated with the progression of diabetic nephropathy on top of baseline albuminuria and chronic kidney disease. We found differences in associations for overall progression and progression from normo- and macroalbuminuria. These novel biomarker discoveries illustrate the utility of analysing urinary metabolites in entire population cohorts.

**Significance Statement:** Individuals with type 1 diabetes are vulnerable to diabetic nephropathy and would benefit from earlier detection of disease progression. Urinary metabolites as a direct read-out of kidney function are potential progression markers. However, analytical tools to quantify a broad panel of urinary metabolites at large scale and low cost are lacking. Recent developments in nuclear magnetic resonance address this need. This study in 2,670 individuals with type 1 diabetes identified ten urinary metabolites associated with progression of diabetic nephropathy. Importantly, different albuminuria categories had different urinary profiles: 2-hydroxyisobutyrate was associated with progression from normoalbuminuria and branched-chain amino acids with progression from macroalbuminuria. These results provide new potential biomarkers and highlight the potential of analysing urinary metabolites on a larger scale.

## Introduction

Type 1 diabetes usually manifests at a young age. Therefore, the affected individuals are at an especially high life-time risk to develop diabetic complications that can substantially lower their quality of life,^1^ shorten their life span^2^ and impose high health costs.^3^ Diabetic nephropathy affects a third of individuals with type 1 diabetes and is the main cause of higher mortality rates in type 1 diabetes.^4^ Although the molecular understanding of diabetic nephropathy is expanding, its exact pathophysiology is still elusive.^5^ Urinary metabolites are a direct readout of kidney function^6^ and might therefore offer novel molecular insights into the pathophysiology of diabetic nephropathy.

Detailed metabolic biomarker profiling, also known as metabolomics, has been successfully used to uncover blood metabolites that are involved at different stages of diabetic nephropathy.^7^ Metabolites in blood are strongly regulated, whereas the metabolite concentrations in urine are more variable and therefore, may offer insights that are not possible from the blood.^8^ Metabolomic profiling of the urine lead for example to the discovery of the role of formate for blood pressure regulation, that had not been discovered in blood.^9^ Urine is abundantly produced by the kidneys and it is easily collected without an invasive procedure. The metabolites found in the urine relate to pathways from cardiometabolic conditions, gut microbial activities to short-term food consumption.^6^ Importantly, urine reflects kidney function and therefore, metabolic biomarker profiling of urine samples is particularly attractive to study kidney disease.^8^ Urinary metabolite studies have identified biomarkers that differentiate between individuals with and without diabetic kidney disease and are linked to mitochondrial metabolism such as 3-hydroxyisovalerate^10,11^ or uraemic solutes that are increased at stage 3 and 4 of chronic kidney disease (CKD).^10^ However, these studies have commonly been limited to case-control studies of modest size^11^ or focused on a small number of metabolites quantified. A lack of analytical tools to robustly quantify a broad panel of urinary metabolites in thousands of individuals (at low cost) has made the regular use of metabolites in large population health studies not feasible. Recently, developments in nuclear magnetic resonance (NMR) have enabled high-throughput metabolomic profiling of urine from large cohorts.^6^

Such recent analytical advances also create enhanced opportunities to identify biomarkers that are related to the progression of diabetic nephropathy. Albuminuria is a strong predictor of progression of diabetic nephropathy, but especially at the early stages, the predictive ability is limited as only around one third of individuals with microalbuminuria experienced a progressive decline in the kidney function during 4 to 10 years of follow-up.^12,13^ In addition, estimated glomerular filtration rate (eGFR), a widely used surrogate marker to assess kidney filtration capacity, is also less suitable at the early stages of diabetic nephropathy due to hyperfiltration^14^ and in general, the prediction of future eGFR is challenging at these early stages.^15^ Therefore, there is need for novel, easily measurable biomarkers that are associated with the progression of diabetic nephropathy.

In this study, we used urine NMR metabolomic profiling in a large cohort of prospectively followed individuals with type 1 diabetes with the aim to identify metabolites associated with the risk of progression of diabetic nephropathy. This knowledge could broaden our understanding of the pathogenesis of diabetic nephropathy and how molecular markers of diet and gut (microbiome) metabolism may reflect disease progression.

## Methods

In this study, we included 2,670 individuals with type 1 diabetes from the Finnish Diabetic Nephropathy (FinnDiane) Study. Type 1 diabetes was defined as age of diabetes onset below 40 years and requiring insulin within the first year of diagnosis. Baseline urinary albumin excretion (AER) was stratified such that normoalbuminuria was defined by an AER below 30 mg/24h, microalbuminuria by an AER of 30 to 300 mg/24h and macroalbuminuria by an AER above 300 mg/24h in two out of three consecutive urine collections. The presence of end-stage kidney disease (ESKD) was defined as being on dialysis or having received a kidney transplant. The five stages of CKD based on eGFR were defined according to the international standard.^16^ At the baseline study visit, the individuals had provided a 24 hours urine sample. We excluded individuals that had no baseline 24-hour urine sample analysed by NMR, and those with no baseline or follow-up information regarding albuminuria. We further excluded individuals with ESKD at baseline and those whose recorded baseline measurement of AER contradicted their characterisation into normoalbuminuria, microalbuminuria or macroalbuminuria at baseline. Individuals were followed for a mean of 9.0 years (standard deviation: 5.0 years) until their first progression event or the latest albuminuria assessment, if they did not progress. Every participant gave written informed consent. The study was approved by the ethics committee of the Helsinki and Uusimaa Hospital District and conducted in accordance with the Helsinki Declaration.

The primary outcome measure was overall progression of diabetic nephropathy defined as an increase in AER to a higher albuminuria category or being diagnosed with ESKD (355 events (13%), 24,145 person-years). In addition, we investigated two subsets: progression from normoalbuminuria (incident diabetic nephropathy) by limiting the participants to those 1,999 individuals with normoalbuminuria at baseline (138 events (7%), 18,655 person-years) and progression from macroalbuminuria by limiting the participants to those 347 individuals with macroalbuminuria at baseline (159 events (46%), 2,613 person-years).

Metabolite quantification of the baseline urine samples was performed using a proprietary NMR metabolite profiling service (Nightingale Health Ltd, Helsinki, Finland). The NMR-based measurements were conducted from 500 µL of stored 24-hour urine samples using a 600 MHz Bruker AVANCE IIIHD NMR spectrometer (Bruker BioSpin, Switzerland) with automated sample changer and cryoprobe. The spectral data were acquired using standard water-suppressed acquisition settings as described in further details in Supplement 1. The urine samples had been stored at -20°C for a median time of 17.8 years (interquartile range: 16.3, 19.7 years) prior to the NMR measurements. The analysis included 54 urinary metabolites quantified in absolute concentration units, as well as the simultaneous quantification of urinary creatinine to enable the analysis of metabolite-to-creatinine ratios. For our main analyses on progression of diabetic nephropathy, three metabolites with over 50% missingness were omitted from analyses in this study (creatine, mannitol, and taurine).

A pre-analytical quality analysis of the influence of storage temperature and storage time on the NMR progression analysis in nine split-aliquote urine samples stored at −20°C and −80°C for a median storage time of 1.8 years did not find any significant differences (Supplement 2). In addition, when comparing creatinine measured by NMR to the one measured by clinical chemistry, with the latter analysed right after the sample collection, we found a strong correlation between both measurements and differences can be largely attributed to storage effects over a median storage time of 17.8 years (Supplement 2).

### Statistics

In a pre-processing step, metabolite concentrations below the detection limit were set to the smallest, detected concentrations for each metabolite. The median and 95% confidence intervals of the absolute baseline concentrations for overall progressors and non-progressors are listed in Table S1. All urinary metabolite concentrations were divided by urinary creatinine to normalise for urine volume (Table S2). All analyses and results apart from Table S1 and the analyses on effect of storage time and temperatures are based on metabolite-to-creatinine ratios, thus the term metabolite refers to its metabolite-to-creatinine ratio, if not otherwise stated explicitly. The distributions of these ratios were skewed for all metabolites; therefore, those ratios were further log-transformed. To facilitate comparison of results for metabolites with different concentration ranges, all hazard ratios are reported per SD-units.

For comparison of baseline clinical characteristics between overall progressors and non-progressors, p-values were calculated with 10,000 permutations (of the progressor/non-progressor labels) and 95% confidence intervals were estimated with bootstrapping and 10,000 iterations. The time-to-event analyses were performed using Cox proportional hazard regression models,^17^ separately for each urine metabolite, with adjustment for sex, calendar year of diabetes diagnosis, baseline age, albuminuria category, CKD stage, and insufficient glycemic control (dichotomous variable: glycated hemoglobin > 58.5 mmol/mol or 7.5%). This (overall) analysis was repeated with a subset of individuals that had normoalbuminuria at baseline and with those with macroalbuminuria.

The proportional hazard assumption was tested with Schoenfeld residuals. To cope with violations of the proportional hazard assumption, all Cox models were stratified according to sex, albuminuria category and CKD stage. If the proportional hazard assumption was violated additionally by the dependent metabolite variable, the follow-up time was stratified into distinct intervals following the method of Zhang et al.^18^ The p-value threshold for significance after adjusting for multiple testing was 0.001. Nominal significance refers to a p-value below 0.05 but above 0.001. Statistical analyses were conducted using R (version 3.6).^19^

## Results

Among the 2,670 individuals with type 1 diabetes included in this study, there were 355 (13%) who progressed to a worse albuminuria category or ESKD, here denoted overall progressors. The baseline characteristics of the progressors differed from the 2,315 non-progressors (Table 1). Progressors were older (39.3 vs. 35.7 years), had a longer duration of diabetes (24.2 vs. 18.5 years) and a higher HbA_1c_ (76.0 vs. 65.0 mmol/mol). Importantly, looking at kidney health, progressors had a lower eGFR (81 vs. 104 ml/min/1.73m^2^) and there were fewer individuals with normoalbuminuria (38.9% vs. 80.4%) but a higher number of individuals with macroalbuminuria (44.8% vs. 8.1%) at baseline. Even though there was no difference in baseline BMI between the two groups, progressors were almost twice as likely to be obese (14.7% vs. 8.0%).

**Table 1.**
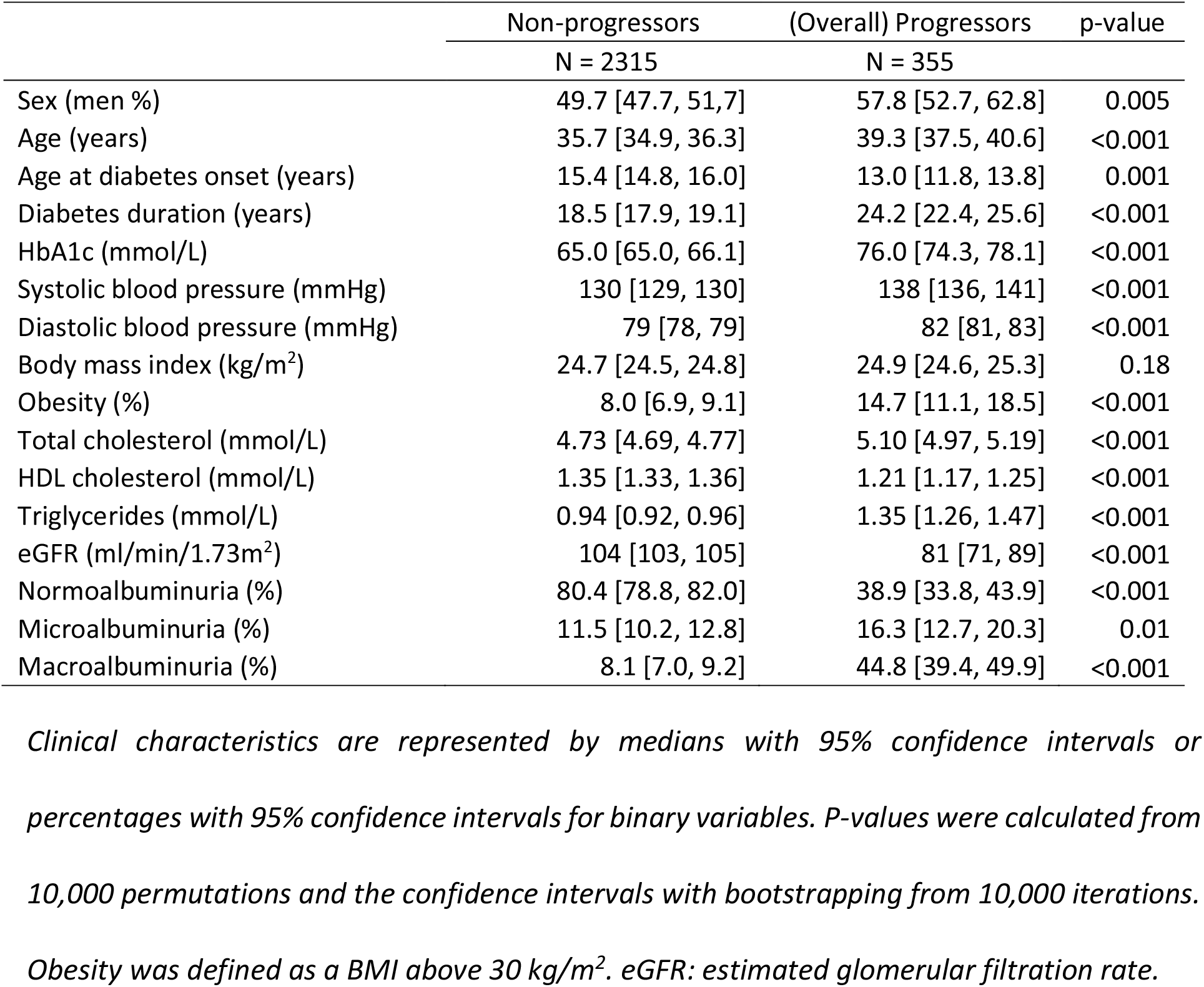
Baseline characteristics of non-progressors and progressors (overall progression).

|  | Non-progressors | (Overall) Progressors | p-value |
| --- | --- | --- | --- |
|  | N = 2315 | N = 355 |  |
| Sex (men %) | 49.7 [47.7, 51.7] | 57.8 [52.7, 62.8] | 0.005 |
| Age (years) | 35.7 [34.9, 36.3] | 39.3 [37.5, 40.6] | <0.001 |
| Age at diabetes onset (years) | 15.4 [14.8, 16.0] | 13.0 [11.8, 13.8] | 0.001 |
| Diabetes duration (years) | 18.5 [17.9, 19.1] | 24.2 [22.4, 25.6] | <0.001 |
| HbA1c (mmol/L) | 65.0 [65.0, 66.1] | 76.0 [74.3, 78.1] | <0.001 |
| Systolic blood pressure (mmHg) | 130 [129, 130] | 138 [136, 141] | <0.001 |
| Diastolic blood pressure (mmHg) | 79 [78, 79] | 82 [81, 83] | <0.001 |
| Body mass index (kg/m<sup>2</sup>) | 24.7 [24.5, 24.8] | 24.9 [24.6, 25.3] | 0.18 |
| Obesity (%) | 8.0 [6.9, 9.1] | 14.7 [11.1, 18.5] | <0.001 |
| Total cholesterol (mmol/L) | 4.73 [4.69, 4.77] | 5.10 [4.97, 5.19] | <0.001 |
| HDL cholesterol (mmol/L) | 1.35 [1.33, 1.36] | 1.21 [1.17, 1.25] | <0.001 |
| Triglycerides (mmol/L) | 0.94 [0.92, 0.96] | 1.35 [1.26, 1.47] | <0.001 |
| eGFR (ml/min/1.73m<sup>2</sup>) | 104 [103, 105] | 81 [71, 89] | <0.001 |
| Normoalbuminuria (%) | 80.4 [78.8, 82.0] | 38.9 [33.8, 43.9] | <0.001 |
| Microalbuminuria (%) | 11.5 [10.2, 12.8] | 16.3 [12.7, 20.3] | 0.01 |
| Macroalbuminuria (%) | 8.1 [7.0, 9.2] | 44.8 [39.4, 49.9] | <0.001 |

*Clinical characteristics are represented by medians with 95% confidence intervals or percentages with 95% confidence intervals for binary variables. P-values were calculated from 10,000 permutations and the confidence intervals with bootstrapping from 10,000 iterations. Obesity was defined as a BMI above 30 kg/m<sup>2</sup>. eGFR: estimated glomerular filtration rate.*

There were significant differences between progressors and non-progressors for 17 metabolites (Table S3). Progressors showed an increased ratio (p<0.0001) for 2-hydroxyisobutyrate, dimethylamine, isoleucine and pseudouridine and a decreased ratio for 3-hydroxyisobutyrate, 3-hydroxyisovalerate, citrate, 4-deoxyerythronic acid, ethanolamine, glycine, glycolic acid, histidine, hypoxanthine, indoxyl sulfate, 1-methylnicotinamide, trans-aconitate and tyrosine.

Assessing the incidence of progression with Cox models, seven urinary metabolites showed a significant association with overall progression after adjustment for sex, age, and diabetes duration as well as baseline glycemic control, albuminuria and CKD stages (Figure 1). For leucine, valine and threonine, the association with progression was significant early on in the follow-up period but did not remain significant after 10 years for leucine and valine or 5 years of follow-up for threonine. Leucine (until 10 years of follow-up) showed the strongest association of all urinary metabolites with overall progression with a hazard ratio (HR) of 1.47 [1.30, 1.66] (p=6.83 × 10^-10^). In comparison, this HR was lower than the one for insufficient glycemic control in individuals with normoalbuminuria (2.41 [1.54, 3.76], p=0.0001) in an equally adjusted model and comparable to the HR for sex (1.34 [0.95, 1.90], p=0.09) from the same model, albeit the latter one was not statistically significant. Valine until 10 years of follow-up (1.38 [1.22, 1.56], p=2.17 × 10^-7^), isoleucine (1.33 [1.18, 1.50], p=5.19 × 10^-6^), 2-hydroxyisobutyrate (1.30 [1.16, 1.45], p=2.40 × 10^-6^), threonine until 5 years of follow-up (1.27 [1.11, 1.46], p=0.0007), and pseudouridine (1.25 [1.11, 1.42], p=0.0002) were also associated with progression of diabetic nephropathy. The relationship was inverse for urinary citrate (0.84 [0.75, 0.93], p=0.0008). The complete results can be found in Figure S5 and Table S4 to support future replication studies and meta-analyses, in accordance with recommended reporting practises for NMR metabolomic profiling studies.^20^

**Figure 1.**
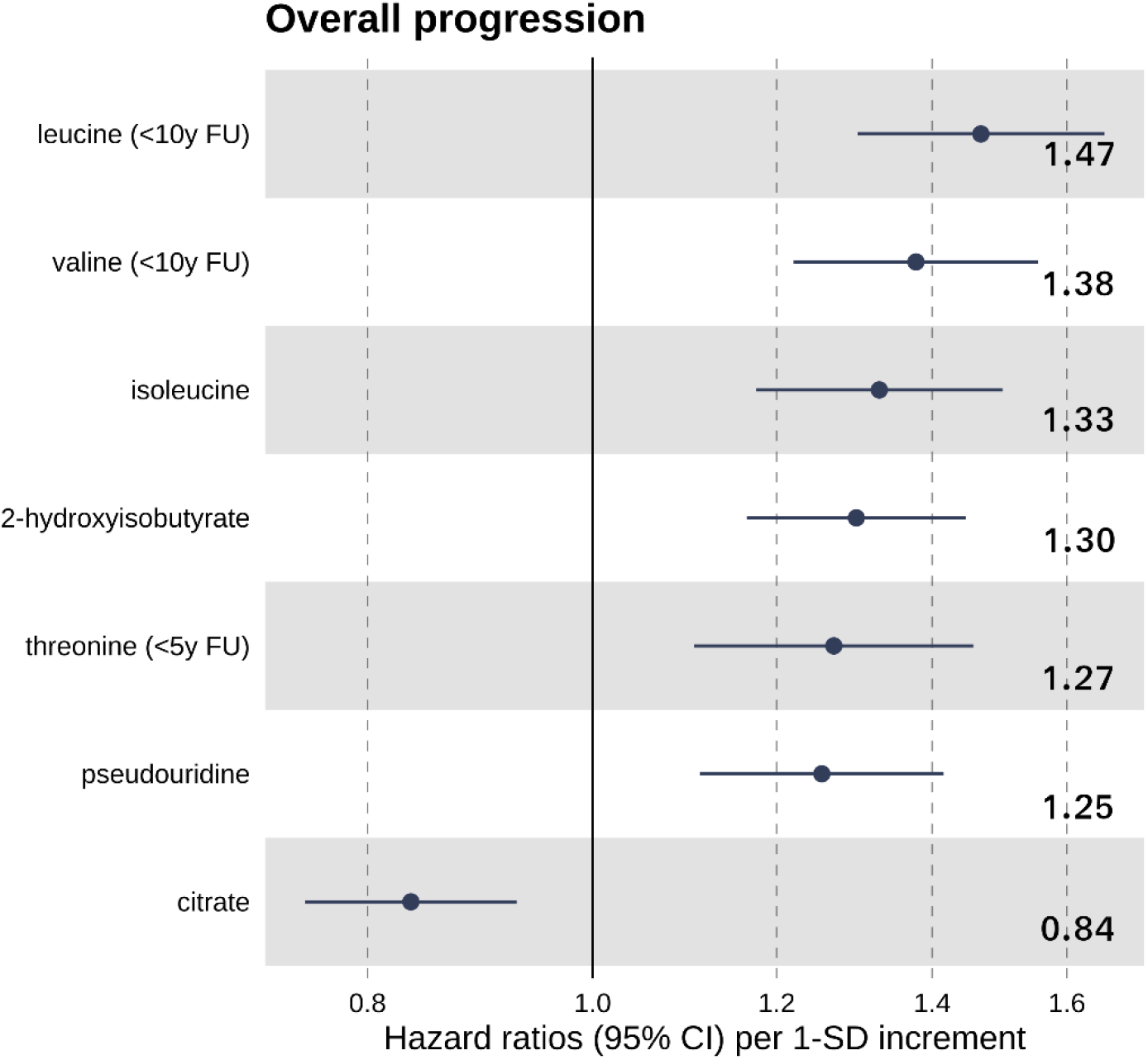
Standardised hazard ratios and 95% confidence intervals for urinary metabolites that were significantly associated with incidence of (overall) progression after accounting for multiple testing (p < 0.001) in all 2,670 individuals. Urine metabolites were scaled to creatinine and log-transformed. The analysis was adjusted for sex and baseline age, year of diabetes diagnosis, baseline glycemic control (HbA_1c_ > 58.5 mmol/mol) and baseline CKD stage and albuminuria class. Hazard ratios were scaled to SD-units. The proportional hazard assumption was tested with Schoenfeld residuals and follow-up times were split when violated. FU: follow-up; y: years.

When looking at the progression from normoalbuminuria only (1,999 individuals, 138 events), 2-hydroxyisobutyrate was significantly associated with incident diabetic nephropathy when adjusted for all covariates including baseline glycemic control, albuminuria and CKD stage with an HR of 1.56 [1.25, 1.95] (p=9.58 × 10^-5^). This metabolite was also associated with overall progression. No other metabolite showed a significant association with progression after taking multiple testing into account (Figure S6, Table S5).

In order to identify urine metabolites that are particularly indicative of progression to the most severe and costly stages of diabetic nephropathy, we further examined associations with the progression from macroalbuminuria to ESKD (347 individuals, 159 events). Six amino acids and one derivate of an amino acid were associated with the incidence of ESKD when adjusted for all covariates including baseline glycemic control, albuminuria and CKD stage (Figure 2). As in the overall results, leucine (1.50 [1.29, 1.73], p=7.50 × 10^-8^), valine (1.42 [1.23, 1.64], p=2.20 × 10^-6^), isoleucine (1.41 [1.19, 1.67], p=7.61 × 10^-5^) and threonine (1.34 [1.18, 1.52], p=6.99 × 10^-6^) were associated with progression. In addition, tyrosine (1.42 [1.20, 1.68], p=4.55 × 10^-5^), alanine (1.32 [1.14, 1.53], p=0.0002) and pyroglutamate (1.37 [1.14, 1.64], p=0.0008) were also associated with incident ESKD. Alanine (up to 5 years of follow-up) was also nominally associated with overall progression (1.24 [1.07, 1.43], p=0.004). The complete results for progression from macroalbuminuria to ESKD can be found in Figure S7 and Table S6.

**Figure 2.**
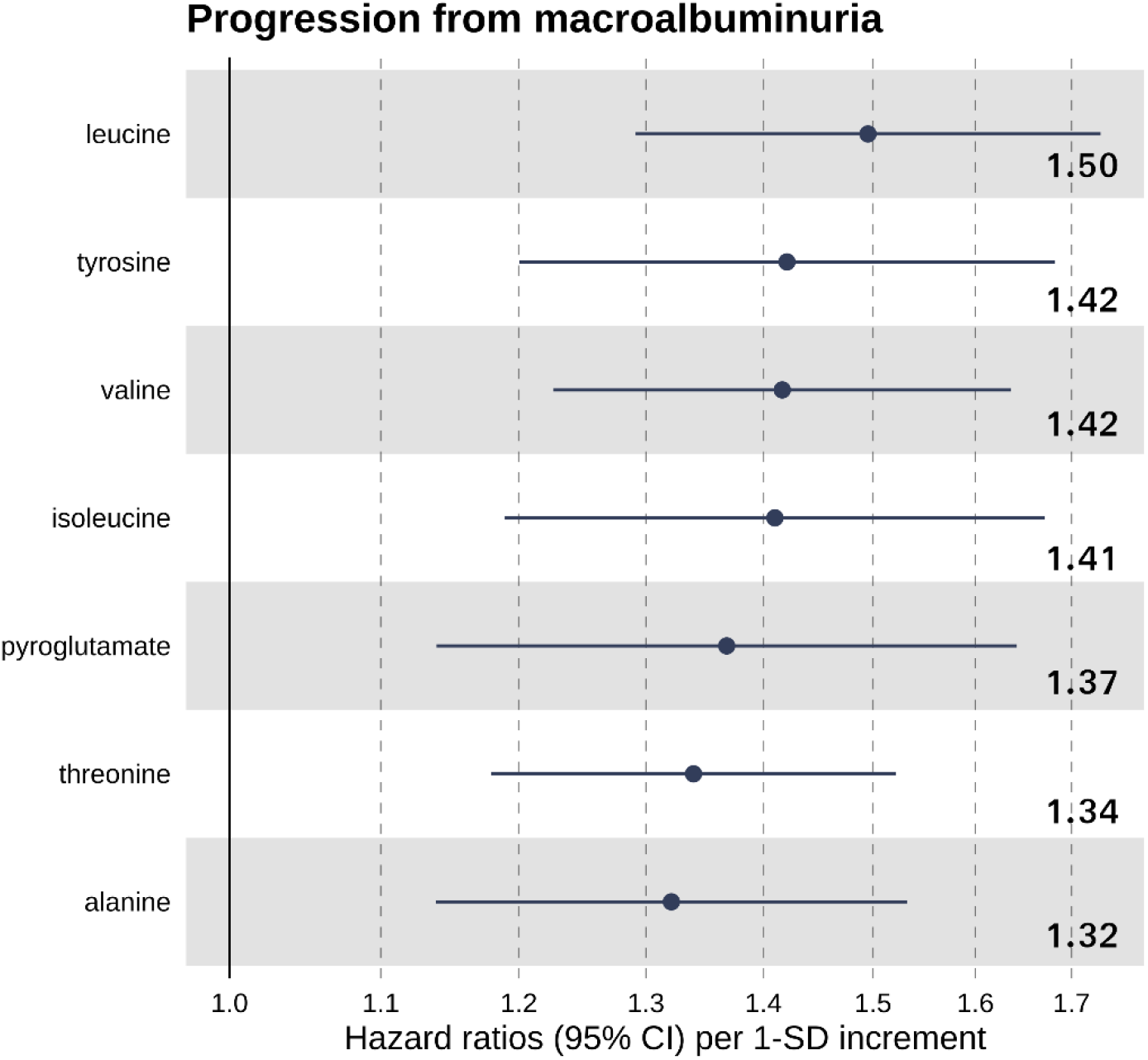
Standardised hazard ratios and 95% confidence intervals for urinary metabolites that were significantly associated with incidence of progression to ESKD after accounting for multiple testing (p < 0.001) in 347 individuals with macroalbuminuria. Urine metabolites were scaled to creatinine and log-transformed. The analysis was adjusted for sex and baseline age, year of diabetes diagnosis, baseline glycemic control (HbA_1c_ > 58.5 mmol/mol) and baseline CKD stage and albuminuria class. Hazard ratios were scaled to SD-units. The proportional hazard assumption was tested with Schoenfeld residuals and follow-up times were split when violated.

Glycine and threonine showed opposing associations when looking at individuals with normoalbuminuria and macroalbuminuria separately. However, most of these associations were only nominally significant. Higher glycine (1.17 [1.02, 1.33], p=0.02) and threonine (1.34 [1.18, 1.52], p=6.99 × 10^-6^) were associated with incident ESKD in those with macroalbuminuria, but lower glycine (0.80 [0.67, 0.95], p=0.01) and threonine (0.81 [0.67, 0.98], p=0.03) were associated with progression from normoalbuminuria (incident diabetic nephropathy).

## Discussion

In this study, we employed a high-throughput NMR platform to analyse 51 metabolite-to-creatinine ratios from 24-hour urine samples of 2,670 individuals with type 1 diabetes and found that 10 ratios were associated with progression of diabetic nephropathy after adjusting for baseline albuminuria category and CKD stage. Importantly, there were differences between the urinary profile for overall progression and progression from normo- and macroalbuminuria suggesting 2-hydroxyisobutyrate as a potential marker for progression from normoalbuminuria (i.e. new-onset diabetic nephropathy) and branched-chain amino acids as markers for overall progression and progression from macroalbuminuria to ESKD.

Several metabolites point towards a link between insulin resistance and progression of diabetic nephropathy. Urinary amino acid, and in particular branched-chain amino acids, were associated with overall progression of diabetic nephropathy as well as progression from macroalbuminuria to ESKD. These novel findings are noteworthy in the context of the many studies on plasma levels of branched-chain amino acids in the field of type 2 diabetes. The first study to show associations between plasma levels of branched-chain amino acids with obesity and insulin resistance dates back 50 years.^21^ These amino acids also represent the most consistent biomarkers identified by blood metabolomics to be predictive of future type 2 diabetes risk.^22–24^ In physiological studies and human genetics, there is even evidence that links them causally to insulin resistance.^25–27^ Furthermore, insulin resistance has been observed in individuals with type 1 diabetes and linked to the increasing prevalence of obesity in type 1 diabetes.^28^ Importantly, in our cohort, progressors were more likely to be obese at baseline (14.73% vs. 7.95%), but linear regressions between the urinary ratios of branched-chain amino acids and waist-height ratio and BMI adjusted for sex, age diabetes onset year, albuminuria category and eGFR showed only a relationship between (central) obesity and urinary isoleucine. However, when calculating the estimated glucose disposal rate (eGDR)^29^ as a surrogate measure for insulin sensitivity and using it in the same regression analysis, all three urinary branched-chain amino acid ratios were associated with lower eGDR values and thus insulin resistance. Interestingly, the correlation for branched-chain amino acids were stronger with glycated hemoglobin than with blood pressure. Therefore, insufficient glycemic control and increasing insulin resistance might be linked to the observed differences in the urine metabolite ratios. Importantly, it has been shown in type 1 diabetes that insulin resistance precedes microalbuminuria.^30^

Urinary 2-hydroxyisobutyrate is another metabolite that links progression of diabetic nephropathy with insulin resistance. It derives from the degeneration of proteins by gut microbiota.^31^ It was shown that urinary 2-hydroxyisobutyrate was higher in obese, insulin-resistant men compared to age-matched, lean controls.^31^ In our cohort, 2-hydroxyisobutyrate was associated with the overall progression, the progression from normoalbuminuria and nominally associated with the progression from macroalbuminuria to ESKD. The fact that 2-hydroxyisobutyrate is associated with both progressions is important as progression from normoalbuminuria is related to changes in albumin excretion and progression from macroalbuminuria is more related to a drop in filtration capacity. In contrast to 2-hydroxyisobutyrate, the branched-chain amino acids did not show any association with progression from normoalbuminuria. Measurements of urine branched-chain amino acids and 2-hydroxyisobutyrate could be important to complement extensive studies on plasma to dissect the role of insulin resistance and the connection to kidney disease progression. It has been shown that insulin resistance precedes microalbuminuria^30^ and furthermore, a small study in individuals with type 1 diabetes found that insulin resistance coincided with a reduction in the glomerular filtration rate.^32^ In our own cohort, we previously found a strong link between albumin excretion rate and insulin resistance.^29^

Urinary pseudouridine has been discussed as a potential glomerular filtration marker and in our study, it was associated with overall progression. Previous studies in serum^33^ have shown elevated concentrations of pseudouridine in renal failure and uremia, and the concentrations were associated with prevalent and incident CKD.^33,34^ Pseudouridine-to-creatinine ratios measured in spot urine were slightly decreased in individuals with CKD, but it is unclear whether these cross-sectional differences were statistically significant.^34^ Previously, pseudouridine was suggested as a glomerular filtration marker, but dismissed due to tubular re-absorption. However, newer findings have shown that pseudouridine and eGFR are correlated and while the calculation of eGFR has to take sex into account, pseudouridine excretion was shown to be independent of sex.^34,35^ Therefore, our results are suggesting that pseudouridine is a glomerular filtration marker.

In addition to filtration, several urinary metabolites are linked to renal tubular damage or protection. We found that low urinary citrate was associated with overall progression of diabetic nephropathy. This observation is in line with a study showing that administration of citrate salts reduces tubulointerstitial injury and slows eGFR decline.^36^ Additionally, metabolic acidosis in individuals with CKD was associated with lower urinary citrate.^37^ In general, our results on citrate are in accordance with a previous small cross-sectional study on CKD^37^ associating urinary citrate with tubular protection.

Previous studies investigating tubular damage showed links with urinary glycine, alanine, and pyroglutamate. In our cohort, we observed differing associations for progression for urinary glycine in individuals with normo- and macroalbuminuria. Lower glycine in urine was nominally associated with progression from normoalbuminuria and higher glycine with progression from macroalbuminuria. Urinary threonine followed the same directions of association and there are several degradation processes for threonine, one leading to the synthesis of glycine.^38^ Low urinary glycine has previously been associated with incident eGFR drop below 60 ml/min/1.73m^2^ in the case-control matched Framingham Offspring cohort of 386 individuals.^39^ This setting is more comparable to our sub-analysis in individuals with normoalbuminuria with a mean eGFR of 105 ml/min/1.73m^2^ at baseline. As in the Framingham Offspring cohort, low urinary glycine was nominally associated with progression from normoalbuminuria. In animal models, dietary glycine has been shown to protect against cyclosporine-mediated proximal tubular damage^39^ and such damage may occur early in kidney disease and might even precede glomerular changes.^40^ We have previously shown that genetic variants in the *GLRA3* gene encoding a glycine receptor are associated with albuminuria.^41,42^ Interestingly, in our cohort, the association changed when looking at the progression from macroalbuminuria. In this scenario, higher and not lower urinary glycine was associated with progression. The reabsorption of glycine through the kidney plays an important role for the bioavailability of glycine.^43^ Low glycine concentrations in serum are linked to a higher incidence of cardiovascular disease with evidence for a causal relationship.^44^ We hypothesise that an increase of glycine in urine is a sign of reduced rate of reabsorption and abnormal kidney function. Along these lines, Nicholson et al. have found increased level of glycine in the urine of rats after inducing damage to the proximal tubule.^45^ They also observed an increase of alanine into the rats’ urine after proximal tubule damage. In our study, increased urinary alanine was associated with incident ESKD in individuals with macroalbuminuria. Urinary pyroglutamate, that was associated with progression from macroalbuminuria to ESKD, has previously been linked to incident macroalbuminuria in individuals with type 2 diabetes and microalbuminuria.^46^ A previous study in type 1 diabetes with 25 individuals linked pyroglutamate to the progression of early kidney disease from normoalbuminuria to microalbuminuria.^47^ However, in our study, we did not find any link between pyroglutamate and such early progression. We suggest that, in a healthier kidney, pyroglutamate is efficiently re-absorbed^48^ and with worsening kidney disease, the reabsorption capabilities are diminishing.

Finally, we also found urinary markers that have been previously linked to responses to oxidative stress and hypoxia such as tyrosine. Oxidative stress is actually a hallmark of CKD^49^ and the interplay between oxidative stress and hypoxia critically contributes to kidney injury.^50^ In our study, higher urinary tyrosine was associated with progression to ESKD in individuals with macroalbuminuria. An inverse association was shown in a previous study in type 2 diabetes that only investigated the earlier stages of kidney disease in individuals that had either normo- or microalbuminuria at baseline.^51^ These observations might be influenced by oxidative stress.^49^ Molnár et al. suggested that both the healthy and the damaged kidney retain para-tyrosine that is converted from phenylalanine. In the presence of free radicals, phenylalanine and consequently tyrosine are hydroxylated in para, meta and ortho positions and the authors observed that the ortho-tyrosine excretion was enhanced in type 2 diabetes through an increased tubular secretion and production.^49^ Therefore, we would indeed expect increased concentrations of urinary tyrosine in the presence of increased oxidative stress.

There are some limitations of our study that need to be acknowledged. The uncovered associations between urine metabolites and the progression of diabetic nephropathy do not allow for any direct causal conclusions. Furthermore, the results have not been replicated in an independent cohort yet as we did not have access to other large cohorts with type 1 diabetes, 24-hour urine collections and long-term follow-up. Large scale urinary NMR analysis, such as this study, are just about to be taken up by many cohorts. In the future, it will probably be possible to validate these results with other large cohorts. The study has also several strengths starting with the size of the cohort. In addition, the urine was measured over 24 hours, which is the gold standard of urine collections. However, the applicability of the results to morning or spot urine collections remains to be addressed. The FinnDiane cohort is thoroughly characterized and has a long follow-up period.

In summary, this study found that 10 out of 51 urinary metabolites were associated with of progression of diabetic nephropathy even after adjusting for baseline albuminuria category and CKD stage. We found differences between overall progression and progression from normo- and macroalbuminuria such as 2-hydroxyisobutyrate as a potential marker for progression from normoalbuminuria. Amino acids and in particular the branched-chain amino acids were strongly associated with progression, and especially with the progression from macroalbuminuria to ESKD. These results provide new potential urinary biomarkers and highlight the potential of routinely analysing urinary metabolites on a larger scale as urinary NMR metabolomic profiling is a reliable high-throughput method.

## Supporting information

EQUATOR Network research reporting checklist

## Data Availability

Due to patient privacy and consent, individual level data is not available for this study. The supplementary tables contain aggregated results per study group.

## Author contributions

P-HG, PW, NS, CF and SM designed the study; VA, KN and LR carried out the NMR metabolite measurements; CF and LMT collected the progression data; EV and SM cleaned, pre-processed and analysed the data; SM drafted the manuscript; P-HG, PW, NS, CF, LMT, KN, EV and SM revised the manuscript; all authors approved the final version.

## Acknowledgements

We thank Hanna Olanne and Heli Krigsman for their skilled laboratory work, selecting the urine samples and sending them for analysis.

