## Supplementary Material for "Urinary metabolite profiling identifies biomarkers for risk of progression of diabetic nephropathy in 2,670 individuals with type 1 diabetes"

#### SUPPLEMENT 1: URINARY METABOLITE PROFILING BY HIGH-THROUGHPUT NMR SPECTROSCOPY

Metabolic profiling of urine in large biobanked sample collections can enable for numerous research applications.[1–4] Here we used established NMR spectroscopy protocols and an automated total-line-shape fitting method to quantify up to 54 urine biomarkers in 24 hours urine samples from 2,670 participants in the prospective FinnDiane study. This method is based on a high-throughput nuclear magnetic resonance metabolomics approach as widely used previously for serum and plasma samples.[5] The automated analyses provide absolute concentrations of 54 urine metabolites and their ratio to creatinine (Supplementary Table S1, S2, S3).

The sample preparation and NMR acquisition parameters were designed for high-throughput rather than optimising for widest possible metabolite coverage. Briefly, urine samples are thawed overnight and subsequently centrifuged (5 min, 3000g). Robotic liquid handlers (JANUS 8-tip workstation; PerkinElmer Inc, USA) are used to mix 70 µL of phosphate buffer (1.5 M $K_2HPO_4$ and 1.5 M $NaH_2PO_4$ in $D_2O$, pH 7.0; including 5.8 mM sodium 3-(trimethylsilyl)propionate-2,2,3,3-$d_4$ and 30.8 mM sodium azide;[6]) and 450 µL urine into 5mm NMR tubes placed on 96-tube racks. For each 96-tube rack, two quality control samples of stock urine are included to enable tracking of consistency of the NMR measurements over time. A 600 MHz Bruker AVANCE IIIHD NMR spectrometer with automated SampleJet sample changer and cryoprobe (CryoProbe Prodigy TCI) is used to acquire the spectral data at 298K. The spectroscopy settings are standard water-suppressed measurements (Bruker noesypresat pulse sequence with mixing time of 10 ms and irradiation field of 25 Hz) using 32 scans per sample with 5.1 s recycle time. A metabolite quantification protocol based on automated implementation of total-line-shape fitting[7] is applied after piece-wise spectral alignment. The scientific literature on chemical shifts and J-couplings for high-abundance metabolites in urine was used to assign metabolite peak shapes.[2,8] The metabolite identifications have further been confirmed with spiking and concentrations calibrated using standard addition.

### SUPPLEMENT 2: STORAGE TEMPERATURE AND STORAGE TIME ANALYSIS

#### Methods

We investigated the analytical effects of the urinary metabolite measures in relation to sample storage temperature and consistency of creatinine measured by clinical chemistry at the time of sample collection. In these pre-analytical quality analysis, split-aliquote urine samples for nine individuals (three women, six men) were stored continuously at both –20˚C and –80˚C for a median storage time of 1.8 years (1.8, 2.4 years). Those nine individuals had a median age of 48.4 years (41.9, 53.0 years) and a median duration of diabetes of 36.8 years (32.4, 43.4 years). We excluded seven biomarkers that were missing for at least five individuals from the full set of 54 markers and assessed the effect of storage time with a linear model with storage time as independent and the logarithm of the ratio of the biomarker's absolute concentrations at −20˚C and −80˚C as dependent variable. In this analysis, a β-coefficient of 0 indicates no effect of storage time. Subsequently, we took advantage of the fact that creatinine was measured clinically at the time the samples were taken and again by NMR for this study. We assessed the association between these two measurements in one linear model for all 2,628 samples that had both measures available and in separate linear models per sample year. The p-value threshold for significance after adjusting for multiple testing was 0.001. Nominal significance refers to a p-value below 0.05 but above 0.001.

#### Results

Storage temperature and storage time did not have a strong influence on the stability of the urine metabolite concentrations over a median follow-up of 1.8 years. When looking at the effect of storing samples at −20˚C and −80˚C, we did not find significant differences after adjusting for multiple testing (Supplementary Table S7). There were only three metabolites where the differences in the absolute concentrations were nominally significant: 4-

deoxythreonate (–0.0023 mmol/L, p = 0.03) and tryptophan (–0.0031 mmol/L, p = 0.03) had slightly lower concentrations at –80˚C compared to –20˚C. Xanthosine concentrations were slightly elevated at – 80˚C compared to – 20˚C (0.012 mmol/L, p = 0.04). We further investigated the Bland-Altman plots (Supplementary Figure S1) to test the agreement of the two measurements. We found two samples (sample 1 and sample 7 in the figure) that were responsible for most of the divergence. The agreement between the two measures was high in general and there was a tendency that the samples diverged in the same direction providing for example higher concentrations at – 20˚C for formate and lower concentrations for xanthosine. When taking storage time into account, we did not find any significant effect after adjusting for multiple testing (Supplementary Table S8). But metabolites such as ethanol (β=0.52, p=0.01), indoxyl sulfate (β=0.15, p=0.02) and especially alanine (β=0.17, p=0.0013) showed a nominal effect of the storage temperature and time. This effect for alanine was likely driven by prominent outliers (Supplementary Figure S2). Notably, we did not observe any significant effect on creatinine (β=0.04, p=0.53). It is important to note that the storage time for this small number of split-aliquote samples was short (median 1.8 years) when compared to the overall storage times of the full set (median 17.8 years).

NMR measurements of urinary creatinine corresponded very well to the urinary creatinine measured by clinical chemistry (β=0.95, $r^2$=0.77) in our study samples (Supplementary Figure S3). In this full set, we found a significant effect of storage time (β=0.02, p=1.9 × 10–13) corresponding to a 1.40% underestimation of creatinine per storage year. The median creatinine measured by clinical chemistry at the time of sample collection was 5.30 mmol/L and 4.34 mmol/L by NMR analysis after a median storage time of 17.8 years (-18%). When we repeated the association analysis only including the latest samples starting from the beginning of 2015, the association between creatinine measured by NMR and clinical chemistry was therefore stronger ($\beta$ = 0.99, $r^2$=0.88). Supplementary Figure S4 summarises the association between creatinine measured by NMR and clinical chemistry by year and highlights that the NMR-measured creatinine quantification reproduced the clinical chemistry measurements of creatinine very well.

#### Conclusions

Urine has been stored at -20˚C prior to the NMR analysis and our quality analysis did not find any significant difference to storage at -80˚C for up to 2 years. When comparing creatinine measured by NMR to the one measured by clinical chemistry, with the latter analysed right after the sample collection, we found a strong correlation between both measurements and differences (-18%) can be largely attributed to storage effects over a median storage time of 17.8 years. As sensitivity analyses, we repeated the main progression analysis with creatinine values adjusted for storage time and additionally with creatinine measured by clinical chemistry. The direction of associations in both sensitivity analyses did not change (data not shown).

### SUPPLEMENT 3: SUPPLEMENTARY TABLE LEGENDS

All supplementary tables are provided separately in Excel format. The table legends are as follows:

Table S1. *Baseline median absolute concentrations and 95% confidence intervals of urinary metabolites for non-progressors and progressors (overall progression). P-values were calculated from 10,000 permutations and the confidence intervals with bootstrapping from 10,000 iterations. A p-value below 0.001 denotes statistical significance.*

Table S2. *Baseline mean metabolite to creatinine ratios and their standard deviations for non-progressors and progressors (overall progression). P-values were calculated from 10,000 permutations and the confidence intervals with bootstrapping from 10,000 iterations. A p-value below 0.001 denotes statistical significance.*

Table S3. *Baseline median metabolite-to-creatinine ratios and 95% confidence intervals for non-progressors and progressors (overall progression). P-values were calculated from 10,000 permutations and the confidence intervals with bootstrapping from 10,000 iterations. A p-value below 0.001 denotes statistical significance.*

Table S4. *Standardised hazard ratios and 95% confidence intervals for all urinary metabolites in the overall Cox regression analysis that assesses progression of diabetic nephropathy for all 2,670 individuals. Urine metabolites were scaled to creatinine and log-transformed. The analysis was adjusted for sex and baseline age, year of diabetes diagnosis, baseline glycemic control ($HbA_{1c}$ > 58.5 mmol/mol) and baseline CKD stage and albuminuria class. Hazard ratios were scaled to SD-units. A p-value below 0.001 denotes statistical significance. The proportional hazard assumption was tested with Schoenfeld residuals and follow-up times were split when violated. FU: follow-up; y: years.*

Table S5. *Standardised hazard ratios and 95% confidence intervals for all urinary metabolites in the Cox regression analysis that assesses progression of diabetic nephropathy for 1,999 individuals with normoalbuminuria at baseline. Urine metabolites were scaled to creatinine and log-transformed. The analysis was adjusted for sex and baseline age, year of diabetes diagnosis, baseline glycemic control (*$HbA_{1c}$ *> 58.5 mmol/mol) and baseline CKD stage and albuminuria class. Hazard ratios were scaled to SD-units. A p-value below 0.001 denotes statistical significance. The proportional hazard assumption was tested with Schoenfeld residuals and follow-up times were split when violated. FU: follow-up; y: years.*

Table S6. *Standardised hazard ratios and 95% confidence intervals for all urinary metabolites in the Cox regression analysis that assesses progression to ESKD for 347 individuals with macroalbuminuria. Urine metabolites were scaled to creatinine and log-transformed. The analysis was adjusted for sex and baseline age, year of diabetes diagnosis, baseline glycemic control (*$HbA_{1c}$ *> 58.5 mmol/mol) and baseline CKD stage and albuminuria class. Hazard ratios were scaled to SD-units. A p-value below 0.001 denotes statistical significance. The proportional hazard assumption was tested with Schoenfeld residuals and follow-up times were split when violated. FU: follow-up; y: years.*

Table S7. *Mean absolute concentration differences and their standard deviations in urine samples for nine individuals stored continuously at –20˚C and –80˚C for a median storage time of 1.8 year. Data from seven metabolites has been removed due to a high number of missing values. P-values were calculated by pairwise t-test. A p-value below 0.001 denotes statistical significance.*

Table S8. *Effect of storage time on the metabolite ratio at –20˚C and –80˚C with a linear model with storage time as independent and logarithm to the base 2 of the ratio as the dependent variable based on split-aliquote urine samples for nine individuals that were stored for a median of 1.8 years. In this analysis, a β-coefficient of 0 indicates no effect of storage time on the ratio. A p-value below 0.001 denotes statistical significance.*

### SUPPLEMENT 4: SUPPLEMENTARY FIGURES

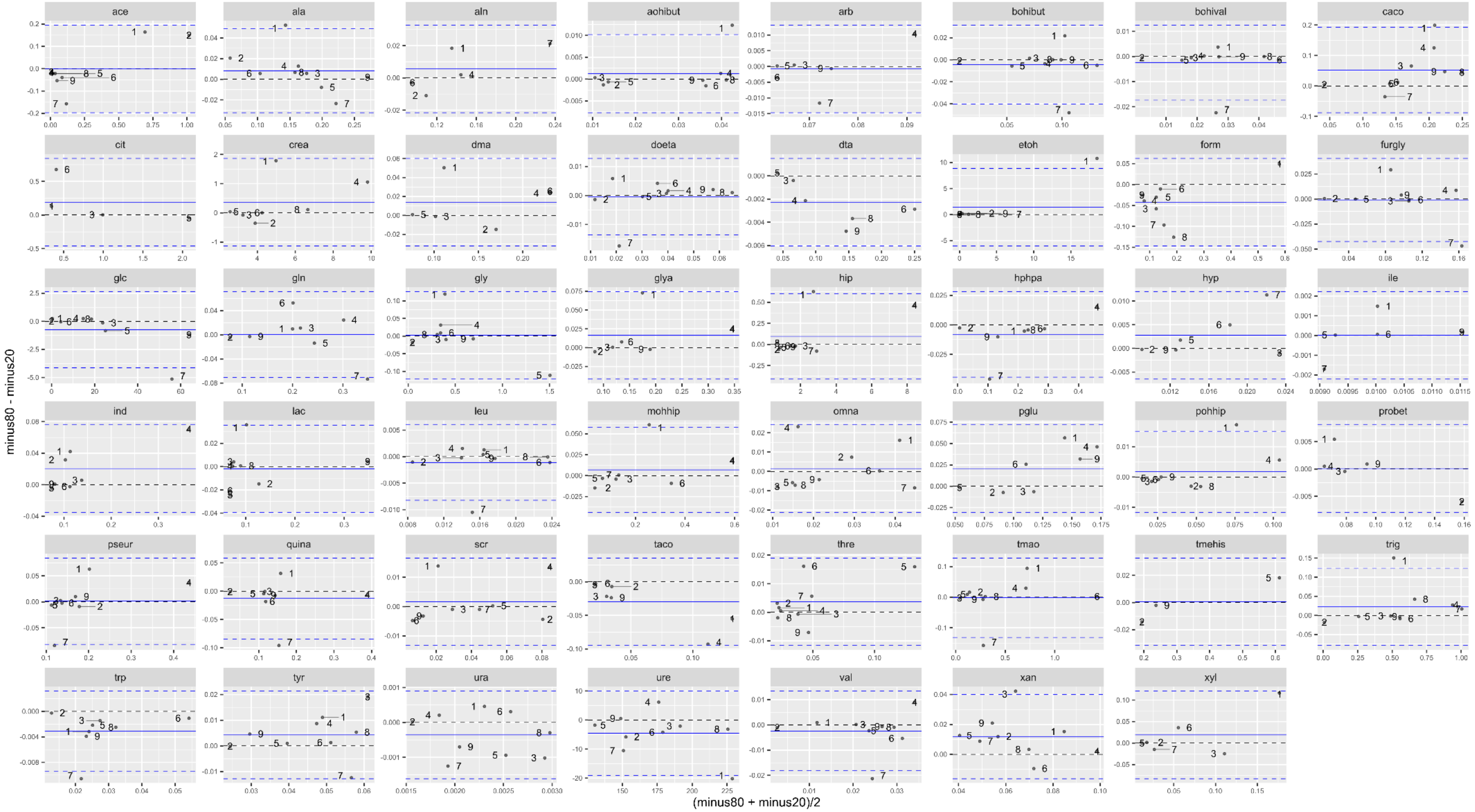

Figure S1. *Bland-Altman plots for nine split-aliquote urine samples from 24h collections stored at –20˚C and –80˚C. The samples are numbered from one to nine and were stored for a median of 1.8 years. The abbreviations for the metabolite names can be found in the list of abbreviations in this supplement.*

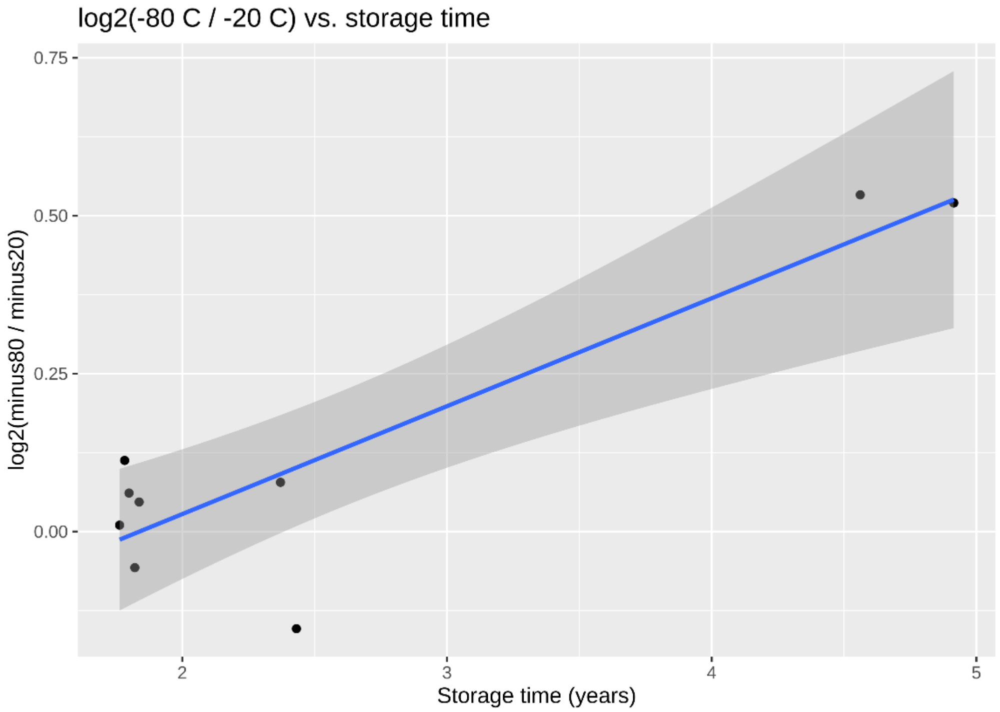

Figure S2. *Relationship of storage time to the logarithm to the base 2 of the metabolite ratio at –20˚C and –80˚C for alanine based on nine split-aliquote urine samples from 24h collections. The samples were stored for a median of 1.8 years.*

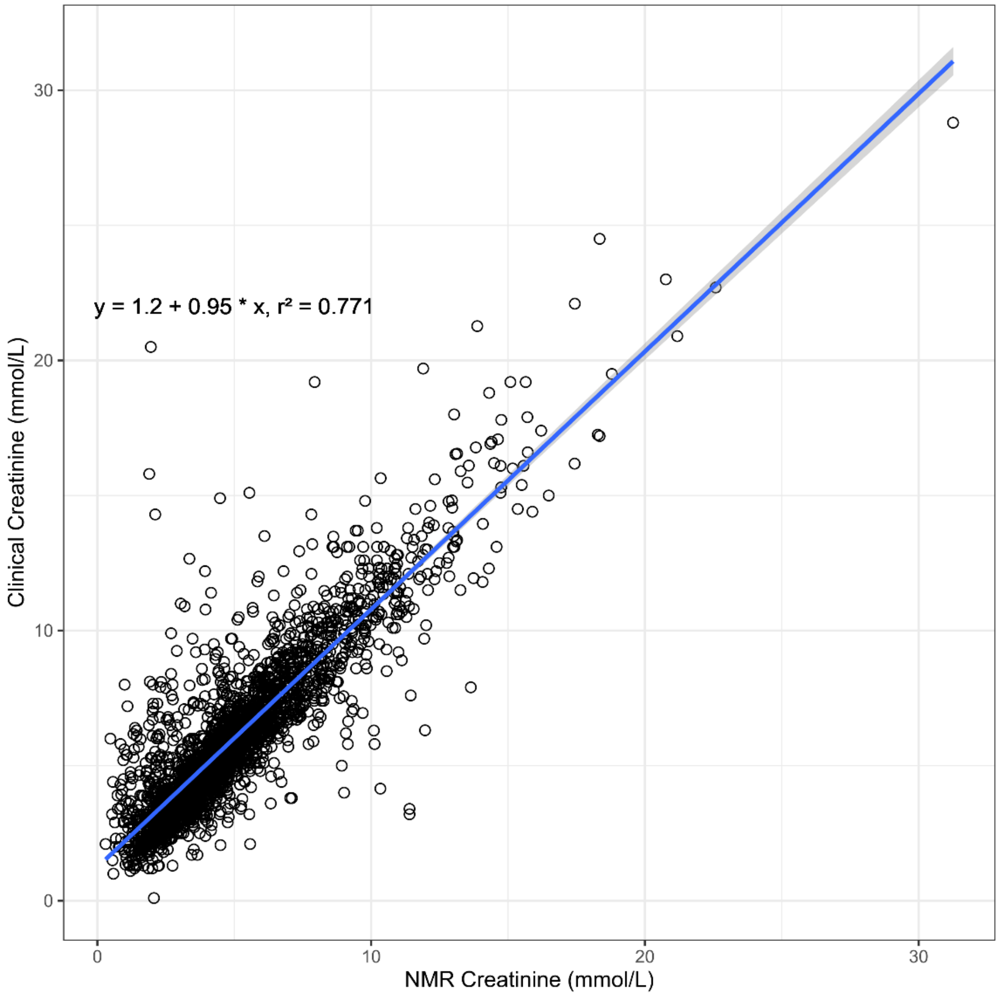

Figure S3. *Correlation of creatinine measured by clinical chemistry at the time of sampling with creatinine measured by NMR after a median storage time of 17.8 years at −20˚C for 2,628 urine samples from 24h urine collections where data was available*

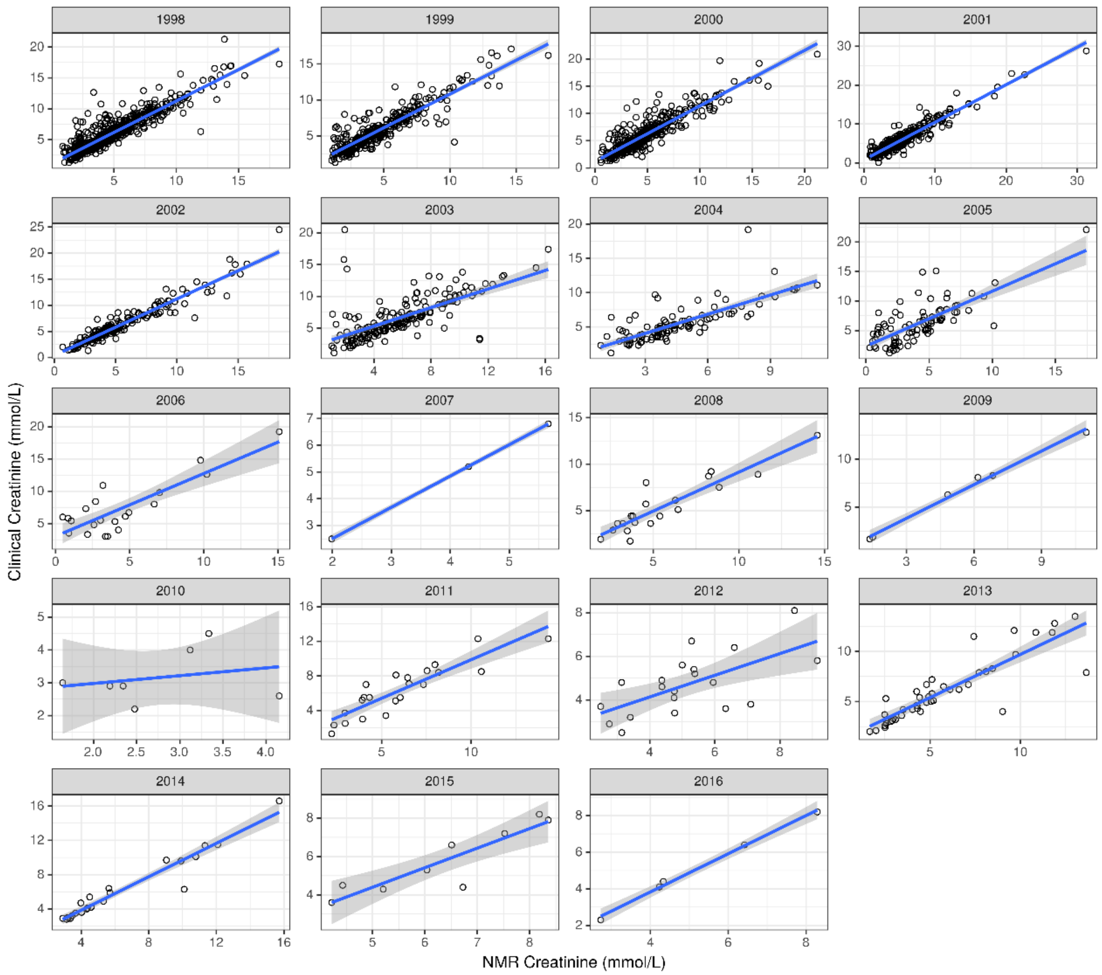

Figure S4. *Correlation of creatinine by clinical chemistry at the time of sampling with creatinine measured by NMR after a median storage time of 17.8 years at –20˚C for 2,628 urine samples from 24h urine collections where data was available grouped by the year of sample taking. The regression lines were fitted with an intercept of 0.*

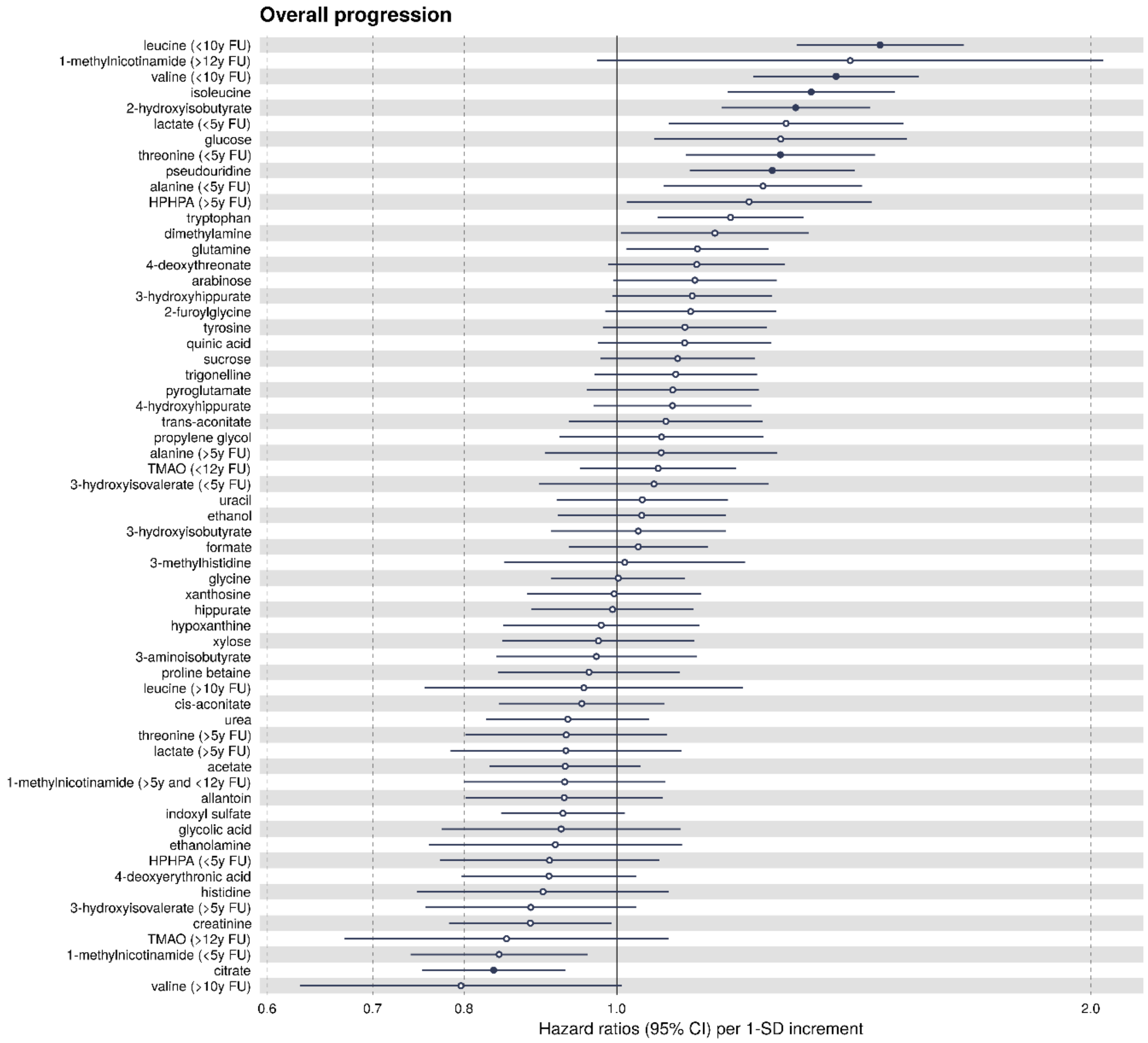

Figure S5. *Standardised hazard ratios and 95% confidence intervals for urinary metabolites associated with (overall) progression of diabetic nephropathy in all 2,670 individuals. Urine metabolites were scaled to creatinine and log-transformed. The analysis was adjusted for sex and baseline age, year of diabetes diagnosis, baseline glycemic control ($HbA_{1c}$ > 58.5 mmol/mol) and baseline CKD stage and albuminuria class. Hazard ratios were scaled to SD-units. A p-value below 0.001 denotes statistical significance and is displayed by a full circle. The proportional hazard assumption was tested with Schoenfeld residuals and follow-up times were split when violated. FU: follow-up; y: years; HPHPA: 3-(3-hydroxyphenyl)-3-hydroxypropionic acid; TMAO: trimethylamine-N-oxide.*

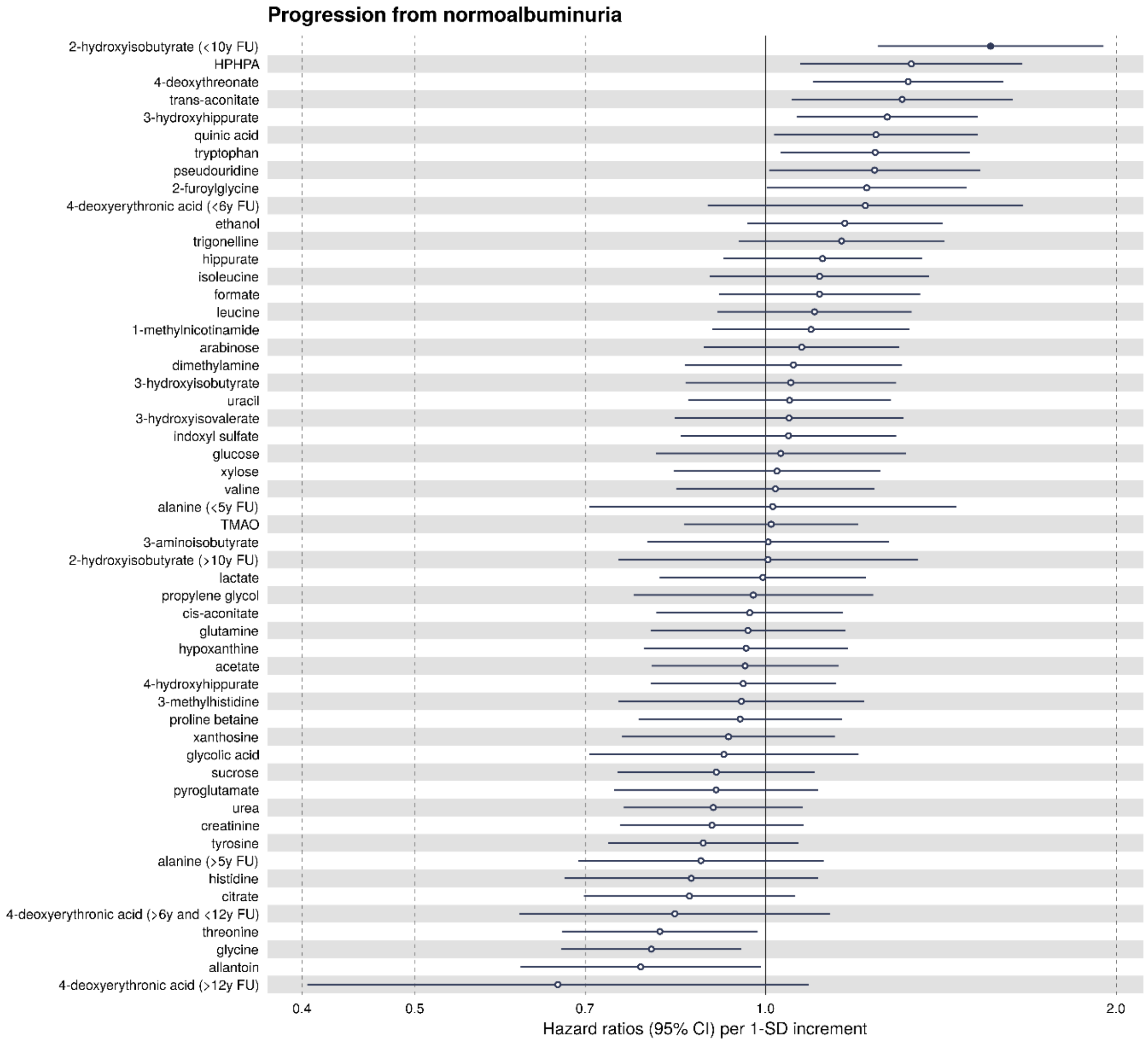

Figure S6. *Standardised hazard ratios and 95% confidence intervals for urinary metabolites associated with progression of diabetic nephropathy in 1,999 individuals with normoalbuminuria at baseline. Urine metabolites were scaled to creatinine and log-transformed. The analysis was adjusted for sex and baseline age, year of diabetes diagnosis, baseline glycemic control ($HbA_{1c}$ > 58.5 mmol/mol) and baseline CKD stage and albuminuria class. Hazard ratios were scaled to SD-units. A p-value below 0.001 denotes statistical significance and is displayed by a full circle. The proportional hazard assumption was tested with Schoenfeld residuals and follow-up times were split when violated. FU: follow-up; y: years; HPHPA: 3-(3-hydroxyphenyl)-3-hydroxypropionic acid; TMAO: trimethylamine-N-oxide.*

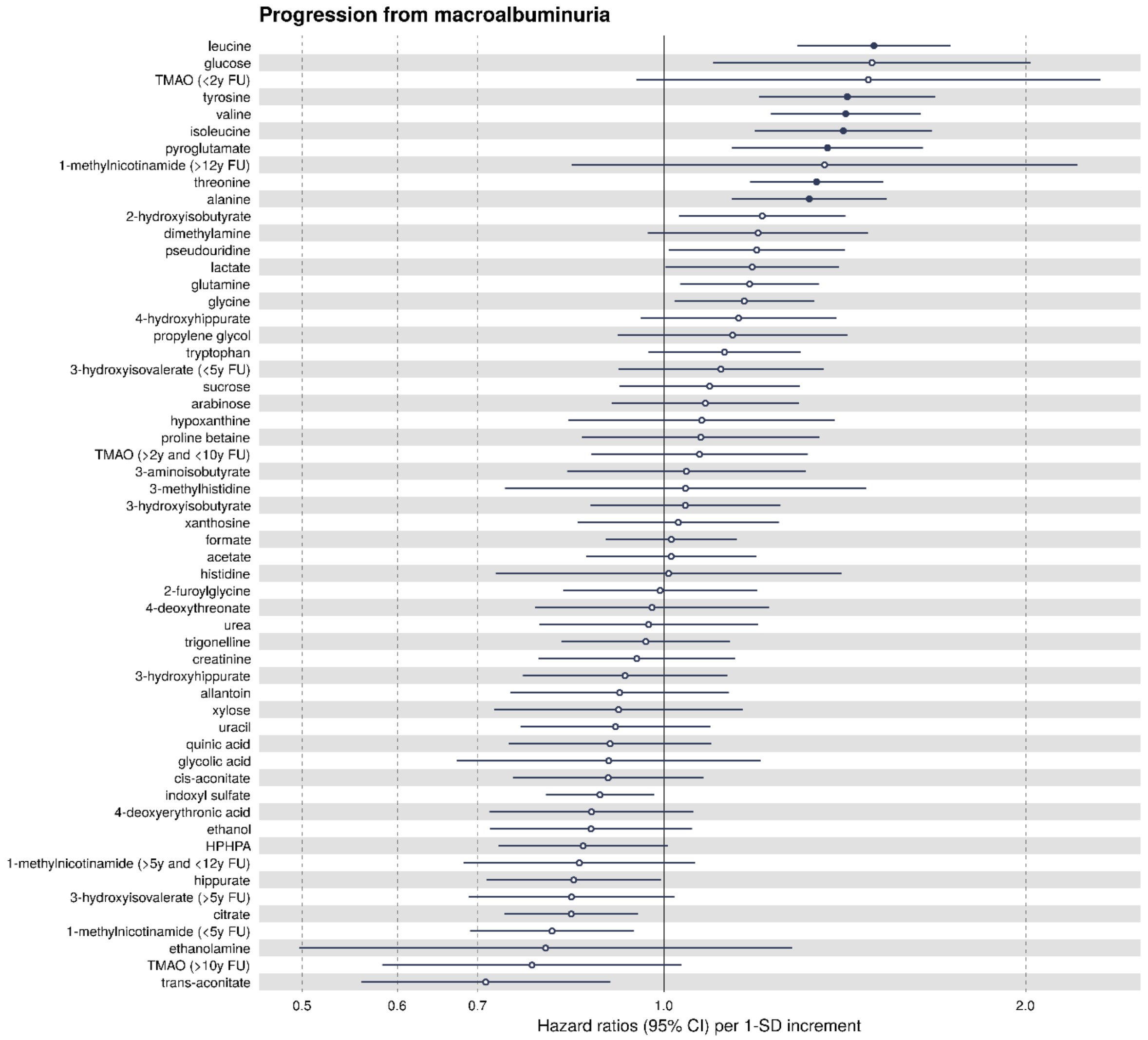

Figure S7. *Standardised hazard ratios and 95% confidence intervals for urinary metabolites associated with incidence of ESKD in 347 individuals with macroalbuminuria at baseline. Urine metabolites were scaled to creatinine and log-transformed. The analysis was adjusted for sex and baseline age, year of diabetes diagnosis, baseline glycemic control ($HbA_{1c}$ > 58.5 mmol/mol) and baseline CKD stage and albuminuria class. Hazard ratios were scaled to SD-units. A p-value below 0.001 denotes statistical significance and is displayed by a full circle. The proportional hazard assumption was tested with Schoenfeld residuals and follow-up times were split when violated. FU: follow-up; y: years; HPHPA: 3-(3-hydroxyphenyl)-3-hydroxypropionic acid; TMAO: trimethylamine-N-oxide.*

#### LIST OF ABBREVIATIONS

Here is a list of abbreviation for metabolite names used in Figure S1:

| | |
|---|---|
| ace | Acetate |
| ala | Alanine |
| aln | Allantoin |
| aohibut | 2-Hydroxyisobutyrate |
| arb | Arabinose |
| bohibut | 3-Hydroxyisobutyrate |
| bohival | 3-Hydroxyisovalerate |
| caco | cis-Aconitate |
| cit | Citrate |
| crea | Creatinine |
| dma | Dimethylamine |
| doeta | 4-Deoxyerythronic acid |
| dta | 4-Deoxythreonate |
| etoh | Ethanol |
| form | Formate |
| furgly | 2-Furoylglycine |
| glc | Glucose |
| gln | Glutamine |
| gly | Glycine |
| glya | Glycolic acid |
| hip | Hippurate |
| hphpa | 3-(3-Hydroxyphenyl)-3-hydroxypropionic acid |
| hyp | Hypoxanthine |
| ile | Isoleucine |
| ind | Indoxyl Sulfate |
| lac | Lactate |
| leu | Leucine |
| mohhip | 3-hydroxyhippurate |
| omna | 1-Methylnicotinamide |
| pglu | Pyroglutamate |
| pohhip | 4-Hydroxyhippurate |
| probet | Proline betaine |
| pseur | Pseudouridine |
| quina | Quinic acid |
| scr | Sucrose |
| taco | trans-Aconitate |
| thre | Threonine |
| tmao | Trimethylamine-N-oxide |
| tmehis | 3-Methylhistidine |
| trig | Trigonelline |
| trp | Tryptophan |
| tyr | Tyrosine |

| | |
|---|---|
| ura | Uracil |
| ure | Urea |
| val | Valine |
| xan | Xanthosine |
| xyl | Xylose |
